# A genomic study of trichotillomania and excoriation disorder in families

**DOI:** 10.1101/2025.09.19.25335566

**Authors:** Samantha R. Greenspun, Isabella Milanes, Luis C Farhat, Sarah Abdallah, Diana Bok, Doris Chen, Wenzhong Liu, Enock Teefe, Michael H Bloch, Thomas V Fernandez, Emily Olfson

**Affiliations:** Child Study Center, Yale School of Medicine, New Haven, CT, USA; Program in translational biomedicine, Yale Graduate School of Arts and Sciences, New Haven, CT, USA; Interdepartmental Neuroscience Program, Yale Graduate School of Arts and Sciences, New Haven, CT, USA; Department of Psychiatry, Faculdade de Medicina FMUSP, Universidade de São Paulo, São Paulo, Brazil; Department of Psychiatry, Yale School of Medicine, New Haven, CT, USA; Wu Tsai Institute, Yale University, New Haven, CT, USA

## Abstract

Trichotillomania and excoriation disorder are obsessive-compulsive related disorders that are often subclassified together as body-focused repetitive behavior (BFRB) disorders. While previous research suggests shared genetic factors, the genetic architecture of these BFRBs remains incompletely understood. Probands with trichotillomania and/or excoriation disorder and both of their biological parents were recruited for an ongoing genetic study of parent-offspring trios with BFRBs. Genome-wide array data were generated in 110 families (334 individuals total) to investigate the role of both common single-nucleotide polymorphisms and rare copy-number variants (CNVs). Polygenic risk scores were calculated using summary statistics from genome-wide association studies of related psychiatric conditions, including obsessive-compulsive disorder (OCD), depression, anxiety, and attention deficit/hyperactivity disorder. Using the polygenic transmission disequilibrium test, we observed a significant over-transmission of polygenic risk for OCD in probands of European ancestry from their parents (mean pTDT = 0.36, *p* = 0.01, n = 92), and a non-significant enrichment for the other conditions. Our results suggest that common variants associated with OCD may contribute to risk for BFRBs, consistent with their current classification as obsessive-compulsive related disorders. We also identified several rare CNVs in probands that overlapped genes intolerant to loss-of-function (LoF) mutations and those previously associated with neurodevelopmental disorders. The LoF-intolerant genes were enriched in biological processes relevant to synapse organization and neurodevelopment. This work provides new insight into the genetic underpinnings of these BFRB disorders, paving the way for larger genomic studies of these understudied conditions.

## Introduction

Trichotillomania (TTM) and excoriation disorder (ED) are obsessive-compulsive related disorders (OCRDs) that impact 1-3% of the population [1, 2, 3, 4]. These body-focused repetitive behavior (BFRB) disorders are characterized by frequent hair pulling or skin picking that results in hair loss or skin damage despite attempts to stop, leading to significant distress and/or functional impairment [5]. These disorders typically onset in early adolescence [6, 7] and are characterized by a waxing-and-waning clinical course [8, 9, 10]. Historically, TTM and ED were thought to be more common in females than males, though recent evidence suggests a more equal gender distribution in the general population [1, 2, 3, 4]. TTM and ED frequently co-occur with other psychiatric conditions, including obsessive-compulsive disorder (OCD), depression, anxiety, and attention deficit/hyperactivity disorder (ADHD) [2, 3]. Currently, these conditions remain difficult to treat, with limited evidence-based treatments available [11, 12, 13, 14]. This highlights a need for a greater understanding of the etiology of TTM and ED, with the hopes that advancements in that knowledge may guide the development of interventions.

Previous family and twin studies suggest that genetic factors contribute to the development of BFRB disorders [15, 16, 17, 18, 19, 20]. In a large twin registry study, Monzani et al. [19] demonstrate that the co-occurrence of OCRDs is best explained by two latent factors, the first of which is common to all OCRDs, while the second is specific to TTM and ED. Despite this role of genetic factors in the etiology of BFRB disorders, there is only one recent genome-wide analysis examining TTM [21] and none of ED. In this recent study, Halvorsen et al. [21] demonstrates a higher polygenic load for generalized psychiatric conditions in 101 individuals with TTM compared to controls, and identifies copy number variants (CNVs) previously associated with other neuropsychiatric disorders, highlighting the value of genomic investigations of BFRB disorders. Given evidence that similar genetic factors contribute risk to both TTM and ED [19], investigating the genetics of TTM and ED together may provide additional insights into the underpinnings of these understudied conditions.

To build on this work, we collected and generated genome-wide array data in 110 families (334 individuals total), consisting of at least one proband with TTM and/or ED and both of their biological parents, making this the first parent-offspring trio genomic study of TTM and ED. This trio design offers advantages over case-control studies by not only enabling the identification of inherited and non-inherited genetic risk but also controlling for potential bias in case-control matching [22]. In these parent-offspring trios, we investigate the transmission of polygenic risk from parents to probands for several commonly co-occurring psychiatric conditions, demonstrating an increased transmission of polygenic risk for OCD in probands with BFRBs. We also identify several rare CNVs that impact genes intolerant to loss-of-function (LoF-intolerant) mutations and neurodevelopmental risk genes that are enriched for canonical processes. These findings provide new insight into the genetics of TTM and ED and lay the groundwork for future larger-scale genomic investigations.

## Methods

### Participants and Procedures

This study involved the recruitment of parent-offspring trios from 110 families in which at least one proband had a diagnosis of TTM and/or ED. Recruitment of 100 of these families was part of an ongoing genetic study (previously described in [23, 24]) approved by the Institutional Review Board (IRB) at Yale School of Medicine (HIC 0301024156). Families were recruited at the TLC Foundation for body-focused repetitive behaviors annual meetings (2018 and 2019) and online through advertisements on the TLC Foundation website, listserv, the study website (tabsstudy.org), and other support groups. The remaining 10 families were recruited as part of a follow-up study of a pediatric TTM clinical trial [25], approved by the IRB at the Yale School of Medicine (HIC 2000022993). Informed consent/assent was obtained from all participants, and each participant was asked to complete a survey about their demographics, co-occurring conditions, family history, and current symptoms. All experimental protocols used in this study follow the standards of the U.S. Federal Policy for the Protection of Human Subjects.

### Genotyping and Sample-level Quality Control

Samples were genotyped using the Illumina Infinium Global Screening Array-24 v3.0 BeadChip at the Yale Keck Biotechnology Resource Laboratory. Quality control was performed using PLINK2 (www.cog-genomics.org/plink/2.0/, Latest Release, 01.29.2025). Individuals were excluded if they had genotype call rates below 98%, high or low levels of heterozygosity (3 standard deviations from the sample mean), or if discrepancies were detected between self-reported gender and sex chromosome data [26]. Relatedness was assessed using KING (https://www.kingrelatedness.com/, Latest Release, 07.28.2024), which uses genetic data to confirm reported relationships [27]. In the presence of admixture, KING accurately infers first-degree (parent-offspring, full siblings, dizygotic twins; kinship coefficient 0.1770–0.3540), second-degree (0.0884–0.1770), and third-degree (0.0442–0.0884) relationships. Any unexpected relationships within or between families were further investigated. Samples with discrepancies in relatedness were removed from the dataset. Overall, 101 probands and their biological parents (289 individuals total) passed these initial quality control steps (details in **Supplementary Figure 2**).

### Imputation and Polygenic Risk Analysis

For polygenic risk score (PRS) analysis, we also restricted the dataset to single-nucleotide polymorphisms (SNPs) that passed additional quality control (QC) filters. SNP-level filtering excluded variants due to missing genotype data, variants that failed Hardy-Weinberg equilibrium testing (*p* < 1 × 10^-6^), and variants with minor allele frequencies (MAF) below 1% using PLINK2 [26]. After QC, 478,854 SNPs remained in the 101 parent-offspring trios for downstream analysis. Imputation was then performed on the QC data using the Sanger Imputation Service against the 1000 Genomes Phase 3 (version 5) reference panel [28]. Post-imputation, we retained 1,204,449 loci with an imputation quality score (Rsq or estimated R²) of 0.3 or higher.

We used GenoPred (opain.github.io/GenoPred) to calculate PRS using the PRS-Continuous Shrinkage (PRS-CS) method [29]. Scores were generated based on genome-wide association studies (GWAS) for OCD [30], major depressive disorder (MDD) [31], anxiety disorders (ANX) [32], and ADHD [33]. All GWAS summary statistics were derived from European ancestry populations and were selected based on being the largest available datasets for each condition at the time of analysis.

PRS were used to perform the polygenic transmission disequilibrium tests (pTDT, https://github.com/ypaialex/ptdt) in the 92 complete parent-offspring trios of European ancestry to assess whether probands inherited a greater polygenic burden for psychiatric disorders than expected by chance. pTDT is calculated by subtracting the mean parental PRS from the proband’s PRS and standardizing this difference by the standard deviation of parental scores [34]. A positive pTDT score suggests that the affected proband inherited a greater genetic risk burden than expected by chance from their parents. To determine whether this deviation from zero was statistically significant, we conducted a two-sided, one-sample t-test.

### CNV calling and quality control

CNVs were called in all BFRB trios that passed general sample-level QC metrics (**Supplementary Figure 2**) with Log R ratio (LRR) and B allele frequency (BAF) data with reference alleles relative to hg38. The calling procedure used PennCNV (http://penncnv.openbioinformatics.org) and cnvPartition (v3.2.1) to generate separate CNV call sets for each sample. CNV loci were validated based on the intersection of these two different call sets, and CNV calls present in both were retained. To ensure high-confidence CNV calls, we excluded poor-quality samples likely to contain spurious CNV calls. Quality control metrics were computed using the PennCNV filter_cnv.pl command and custom in-house scripts. Outlier detection focused on four sample-level metrics: Log R Ratio standard deviation (LRR_SD), absolute waviness factor (|WF|), BAF drift, and number of CNV calls (numcnv). Samples exceeding any of the following thresholds were removed: LRR_SD > 0.3, |WF| > 0.05, BAF drift > 0.01, or numcnv ≥ 20. This resulted in a further exclusion of 15 trios from the initial 101 trios that passed QC, with the final set for CNV analysis comprising 86 trios (**Supplementary Figure 2**). After applying these filters, all retained samples met the specified quality control criteria. Additional details on PennCNV and cnvPartition are in the Supplemental Methods.

### CNV filtering and annotation

CNV calls underwent several filtering steps to generate a final set of rare CNVs (MAF < 1%) for downstream analyses, as done in previous studies [35, 36]. First, we removed CNVs that overlapped genomic regions (defined as > 50% overlap), known to harbor spurious CNV calls: immunoglobulin regions, segmental duplications, or loci within 500 kb of telomeres or centromeres. We also removed CNVs where 50% of bases overlapped loci from Repeatmasker (version 4.0.5, http://www.repeatmasker.org) or “non-defined” (i.e., polyN) portions of the hg38 reference panel. CNVs that were adjacent were merged into one single call if they overlapped by ≥ 30% of the total base-pair length. Calls were then subjected to a series of CNV size and frequency filters. Specifically, calls were required to be greater than 30 kb in size, found in less than 1% of the population in both gnomAD (version 4.1.0) structural variants global and European datasets (covering over 14,000 individuals), and in our CNV dataset. CNVs were also required to have a confidence level greater than 50, as done previously [37, 38]. We also examined rates of CNV calls that were both rare (MAF < 1%) and large (≥ 500 kb).

Classification of CNVs as either *de novo* or transmitted was based on parental CNVs and was confirmed by visualizing both the offspring’s and parents’ CNV profiles. See the Supplemental Methods section for a description of the full procedure.

Rare CNVs that passed filtering criteria were annotated to genes using the UCSC hg38 database. CNV calls were categorized as deletions if copy number ≤ 1 or duplications if copy number ≥ 3. Genes overlapping with CNVs were then assessed for loss-of-function (LoF) intolerance, defined as a pLI score ≥ 0.9 in gnomAD (version 4.1.0). These genes were also checked to see if they have been reported in other neuropsychiatric and neurodevelopmental conditions. This included CNVs previously associated with neurodevelopmental disorders defined by Kendall et al. [39], and CNVs predicted to disrupt neurodevelopmental risk genes defined by an FDR ≤ 0.05 in Fu et al. [40]. These categories were chosen to match a recent large case-control CNV study in OCD [35] to enable comparison.

### Exploratory gene-ontology analysis

We used ConsensusPathDB (http://cpdb.molgen.mpg.de/, Latest Release 35, 05.06.2021) to evaluate whether the set of LoF-intolerant genes that were predicted to be disrupted by a rare CNV (**Tables 2 and 3**) was significantly over-represented in 31 public gene ontology sets. **Supplementary Table 2** provides the p-values (< 0.01) and q-values (< 0.05) for all enriched gene ontology sets. Additional details on how this data was generated can be found in the Supplementary Methods.

## Results

### Demographic and clinical characteristics

After quality control, we examined 101 probands with a diagnosis of TTM, ED, or both conditions and their biological parents (289 individuals total, including 87 parent-offspring trios and 7 quads with two affected probands). The mean age at assessment for probands was 22 years, and the cohort was predominantly female (94%) (**Table 1**). Based on the principal component analysis, most probands were of European ancestry (n = 95), with smaller representation from Admixed American (n = 4), East Asian ancestry (n = 1), and Central and South Asian ancestry (n = 1) (**Table 1**). Trios were classified as simplex if the proband had no first-degree relative with a history of trichotillomania or excoriation disorder (n = 41), and as multiplex if at least one first-degree relative had a history of one of these BFRBs (n = 58) (**Table 1**).

**Table 1.**
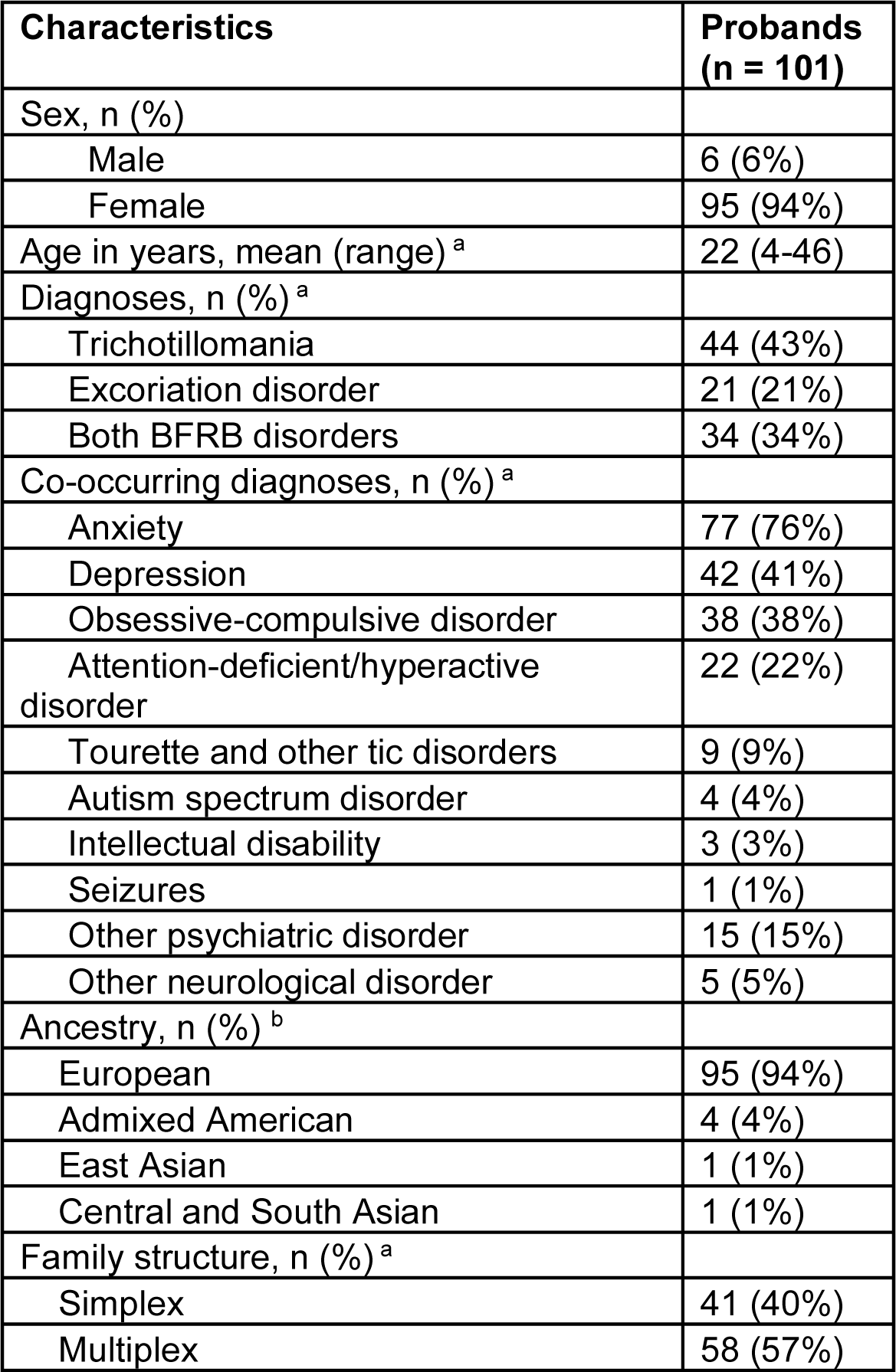
Demographic and clinical characteristics of probands with trichotillomania and/or excoriation disorder. Characteristics of 101 probands with trichotillomania and/or excoriation disorder who passed initial quality control of genotyping data and are included in the genomic analyses. ^a^Two probands who completed consents and reported TTM and/or ED at the time of recruitment, but did not complete the surveys, so their age, diagnoses, and family structure are not included in this table. ^b^Ancestry was determined by principal components analysis.

| Characteristics | Probands<br>(n = 101) |
| --- | --- |
| Sex, n (%) |  |
| Male | 6 (6%) |
| Female | 95 (94%) |
| Age in years, mean (range)<sup>a</sup> | 22 (4-46) |
| Diagnoses, n (%)<sup>a</sup> |  |
| Trichotillomania | 44 (43%) |
| Excoriation disorder | 21 (21%) |
| Both BFRB disorders | 34 (34%) |
| Co-occurring diagnoses, n (%)<sup>a</sup> |  |
| Anxiety | 77 (76%) |
| Depression | 42 (41%) |
| Obsessive-compulsive disorder | 38 (38%) |
| Attention-deficient/hyperactive disorder | 22 (22%) |
| Tourette and other tic disorders | 9 (9%) |
| Autism spectrum disorder | 4 (4%) |
| Intellectual disability | 3 (3%) |
| Seizures | 1 (1%) |
| Other psychiatric disorder | 15 (15%) |
| Other neurological disorder | 5 (5%) |
| Ancestry, n (%)<sup>b</sup> |  |
| European | 95 (94%) |
| Admixed American | 4 (4%) |
| East Asian | 1 (1%) |
| Central and South Asian | 1 (1%) |
| Family structure, n (%)<sup>a</sup> |  |
| Simplex | 41 (40%) |
| Multiplex | 58 (57%) |

### Polygenic transmission for psychiatric disorders

Results demonstrate a significant over-transmission of OCD polygenic risk in probands with BFRBs from their parents (mean pTDT = 0.36, *p* = 0.01) (**Figure 1.A**). For depression, anxiety, and ADHD, transmission of polygenic risk from parents to probands with BFRBs was not statistically enriched, though in the positive direction (**Figure 1.A**). We performed a sensitivity stratified analysis of probands who did and did not self-report a co-occurring OCD diagnosis (**Figure 1.B)**, demonstrating a positive, non-significant transmission of OCD polygenic risk in both groups (mean pTDT_No co-occurring OCD_ = 0.35, *p* = 0.06, n = 57; mean pTDT_Co-occurring OCD_ = 0.37, *p* = 0.06, n = 35) with no statistically significant difference between these two groups (*p* = 0.94).

**Figure 1.**
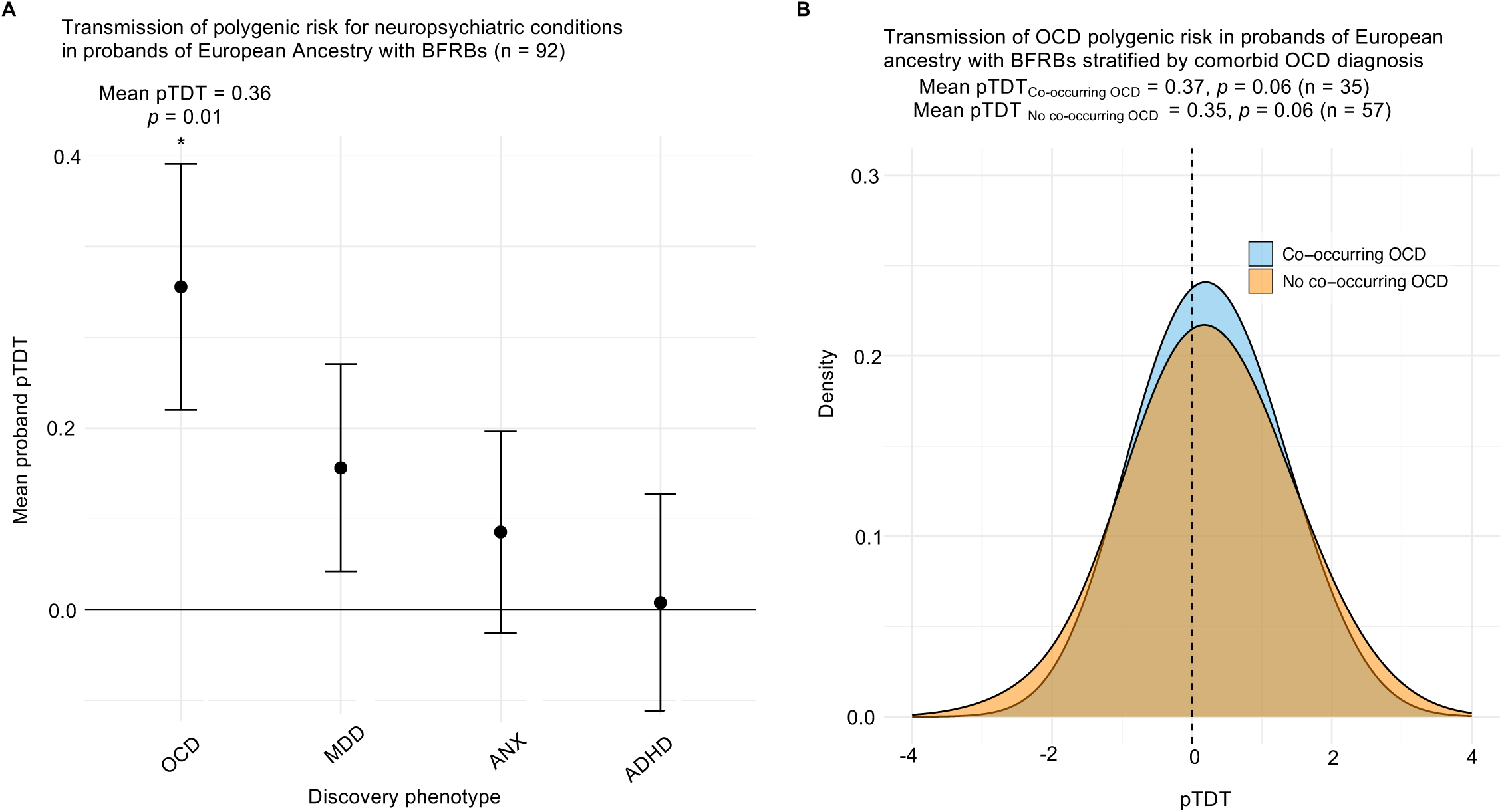
Transmission of polygenic risk for psychiatric conditions in probands of European Ancestry with trichotillomania and/or excoriation disorder. A) Transmission of polygenic risk for several related psychiatric conditions in probands with body-focused repetitive behavior disorders (BFRBs). The polygenic transmission disequilibrium test (pTDT) compares the mean deviation of proband polygenic risk scores to the mid-parent distribution. A significantly increased transmission of polygenic risk for obsessive-compulsive disorder (OCD) was observed in the probands with BFRBs (mean pTDT = 0.36, *p* = 0.01). Transmission of polygenic risk was not statistically significant for other neuropsychiatric conditions (MDD pTDT = 0.15, *p* = 0.17; ANX pTDT = 0.08, *p* = 0.44; ADHD pTDT = 0.01, *p* = 0.94). Error bars indicate standard errors. ∗*p* < .05 in a two-sided, one-sample t-test. B) Sensitivity analysis examining transmission of polygenic risk for OCD in BFRB probands with or without a self-reported co-occurring OCD diagnosis. Probands with BFRBs who have an OCD diagnosis and those who do not show a mean pTDT in the positive direction, but it was not statistically significant in either group (mean pTDT_No co-occurring OCD_ = 0.35, *p* = 0.06, n = 57; mean pTDT_Co-occurring OCD_ = 0.37, *p* = 0.06, n = 35). The difference between the groups was not statistically significant (*p* = 0.94). OCD, obsessive-compulsive disorder; MDD, major depressive disorder; ANX, anxiety disorders; ADHD, attention-deficient/hyperactivity disorder.

To further understand patterns of OCD polygenic risk transmission in these families with BFRBs, we also conducted post-hoc exploratory secondary analyses to investigate the impact of BFRB disorder diagnosis (TTM, ED, vs both) and family structure (simplex vs multiplex) (**Supplementary Figure 3.A and Supplementary Figure 3.B**). For BFRB diagnosis, the mean pTDT for OCD polygenic risk was in the positive direction for all three BFRB disorder diagnosis groups, but was only statistically significant for those with a singular diagnosis of TTM (mean pTDT = 0.56, *p* = 0.01) (**Supplementary Figure 3.A**). Probands from simplex families showed a significant over-transmission of OCD polygenic risk (mean pTDT = 0.67, *p* = 0.001), but not those from multiplex families (mean pTDT = 0.15, *p* = 0.4) (**Supplementary Figure 3.B**). In these exploratory secondary analyses, the differences in mean pTDT were not statistically significant between groups.

### Copy-number variant analysis

We assessed rates of rare (MAF < 1%) CNVs in probands with TTM and/or ED, showing an overall rate of 0.73 rare CNVs per proband with a deletion rate of 0.32 and a duplication rate of 0.40 (**Table 2**). No rare *de novo* CNVs were identified after visualization of the Log R Ratios and B allele frequencies, so our downstream analyses focused on rare transmitted CNVs. In addition to rare CNVs, we also examined the rate of large (length ≥ 500 kb), rare CNVs in BFRB probands, with a deletion rate of 0.01 and a duplication rate of 0.03.

**Table 2.**
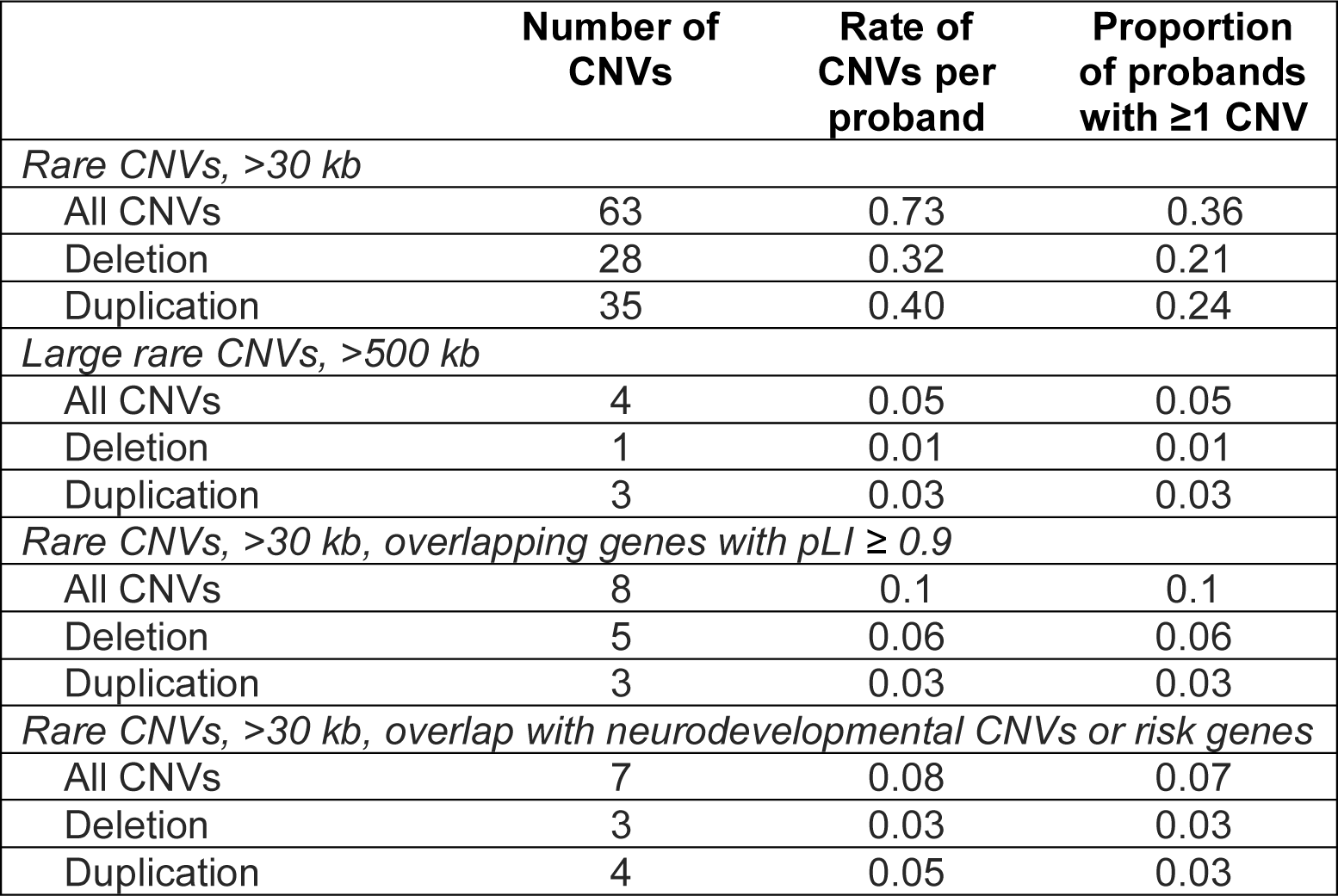
Rates of rare copy number variants in probands with trichotillomania and/or excoriation disorder. Rates of different categories of rare copy number variants (CNVs) identified by both PennCNV and cnvPartition in 86 probands with trichotillomania and/or excoriation disorder who passed quality control steps for CNV analyses. Rare CNVs were defined as <1% in gnomAD (version 4.1.0) structural variants in the global, European datasets, and in our CNV dataset. Large CNVs were defined as >500 kb. Rare copy number variants (CNVs) impacting genes with high loss-of-function intolerance were defined based on protein loss-of-function intolerance (pLI) scores ≥ 0.9. CNVs previously associated with neurodevelopmental disorders were defined by Kendall et al. [39], and CNVs harboring neurodevelopmental risk genes with an FDR < 0.05 in Fu et al. [40]. kb, kilobases.

|  | **Number of CNVs** | **Rate of CNVs per proband** | **Proportion of probands with $\geq 1$ CNV** |
| --- | --- | --- | --- |
| *Rare CNVs, >30 kb* |  |  |  |
| All CNVs | 63 | 0.73 | 0.36 |
| Deletion | 28 | 0.32 | 0.21 |
| Duplication | 35 | 0.40 | 0.24 |
| *Large rare CNVs, >500 kb* |  |  |  |
| All CNVs | 4 | 0.05 | 0.05 |
| Deletion | 1 | 0.01 | 0.01 |
| Duplication | 3 | 0.03 | 0.03 |
| *Rare CNVs, >30 kb, overlapping genes with $pLI \geq 0.9$* |  |  |  |
| All CNVs | 8 | 0.1 | 0.1 |
| Deletion | 5 | 0.06 | 0.06 |
| Duplication | 3 | 0.03 | 0.03 |
| *Rare CNVs, >30 kb, overlap with neurodevelopmental CNVs or risk genes* |  |  |  |
| All CNVs | 7 | 0.08 | 0.07 |
| Deletion | 3 | 0.03 | 0.03 |
| Duplication | 4 | 0.05 | 0.03 |

Using the set of genes that overlapped rare CNVs identified in probands, we investigated whether these genes showed high intolerance to LoF variation or were previously associated with neurodevelopmental risk. Eight rare CNVs from eight probands overlapped genes with high LoF intolerance scores (pLI ≥ 0.9) (**Table 3**). In addition, we identified five CNVs previously associated with neurodevelopmental disorders by Kendall et al. [39], including deletions in *NRXN1*, 10q23, and *PWS/AS*, and duplications in *WBS* and 8p23.1 (**Table 3** and **Supplementary Table 1.1**). We further mapped these CNVs to the specific genes associated with the neurodevelopmental disorder (*NRXN1*, *FKBP6*, *TRIM50*, *PTEN*, and *PWRN1)*. Four rare CNVs overlapped high-confidence risk genes associated (FDR < 0.05) with neurodevelopmental disorders (NDDs) in Fu et al. (2022) [40], including *KIF1A, NRXN1, NIPBL*, and *PTEN* (**Table 3** and **Supplementary Table 1.2**). Visual representations of these individual CNVs and neurodevelopmental FDR results are available in the supplement (**Supplementary Figure 4** and **Supplementary Table 1.2**, respectively). Characteristics of individuals with these rare CNVs impacting LoF-intolerant and neurodevelopmental genes are detailed in **Table 3**, and it is notable that none of these probands self-reported a history of autism or intellectual disability.

**Table 3.** Rare copy number variants identified in probands with BFRBs that overlap with LoF-intolerant genes, neurodevelopmental CNVs, and neurodevelopmental risk genes. Rare copy number variants (CNVs) found in 10 probands with BFRBs that overlap loss-of-function (LoF) intolerant genes (pLI ≥ 0.9), known neurodevelopmental CNVs, and neurodevelopmental risk genes (indicated by boldness). CNVs previously associated with neurodevelopmental disorders were defined by Kendall et al. [39], and CNVs harboring neurodevelopmental risk genes with an FDR ≤ 0.05 in Fu et al. [40]. TTM, trichotillomania; ED, excoriation disorder; OCD, obsessive-compulsive disorder; TD, tic disorder; ANX, anxiety disorder; MDD, major depressive disorder; ADHD, attention-deficient/hyperactivity disorder; DEL, deletion; DUP, duplication; WBS, Williams-Beuren syndrome; PWS/AS, Prader-Willi Syndrome/Angelman Syndrome.

| ID | Sex | BFRB diagnosis | Co-occurring conditions | Genomic coordinates (hg38) | CNV class | Size, bp | Known CNV | Gene(s) of interest impacted (pLI score if $\geq 0.9$) | Other known genes impacted |
| --- | --- | --- | --- | --- | --- | --- | --- | --- | --- |
| BFRB9003.p1 | Female | ED | OCD, ANX | 2:240684041-240762707 | DUP | 78,667 | - | **KIF1A** (1) | **AQP12A** |
|  |  |  |  | 8:73040513-73159092 | DUP | 118,580 | - | **TERF1** (0.9) | **SBSPON** |
| BFRB9021.p1 | Female | TTM | MDD, ANX | 2:50652887-50725225 | DEL | 72,339 | NRXN1 del | **NRXN1** (1) | **MIR8485** |
| BFRB9023.p1 | Female | TTM | OCD, ANX | 7:73308984-73391588 | DUP | 82,605 | WBS dup | **FKBP6**, **TRIM5** | **FKBP6**, **TRIM5**, **RNU6-1080P**, **RPL7AP77** |
| BFRB9047.p1 | Female | TTM | OCD, MDD, ANX | 5:37000986-37046181 | DEL | 45,196 | - | **NIPBL** (1) | - |
|  |  |  |  | 10:87894024-87925562 | DEL | 31,539 | 10q23 del | **PTEN** | - |
| BFRB9059.p1 | Female | TTM | ANX | 9:101144710-101244151 | DEL | 99,442 | - | **PLPPR1** (0.94) | - |
| BFRB9061.p1 | Female | TTM | ADHD, ANX | 15:24140203-24249920 | DEL | 109,718 | PWS/AS del | **PWRN1** | **PWRN2** |
| BFRB9072.p1 | Female | ED | Other BFRB, ANX | 8:8243957-8465532 | DUP | 221,576 | 8p23.1 dup | **PRAG1** | **FAM86B3P**, **LINC02949**, **LINC02950** |
| BFRB9082.p1 | Female | TTM | MDD, ANX | 14:19202105-19948340 | DUP | 746,236 | - | **POTEG** (1) | **ARHGAP42P4**, **BMS1P17**, **CDRT15P13**, **DUXAP10**, **GRAMD4P3**, **LINC01297**, **MED15P6**, **NBEAP6**, **NF1P11**, **NF1P4**, **OR11H2**, **OR11K2P**, **OR4H12P**, **OR4K1**, **OR4K2**, **OR4K3**, **OR4K4P**, **OR4K5**, **OR4K6P**, **OR4M1**, **OR4N1P**, **OR4N2**, **OR4Q3**, **RNU6-1268P**, **TOMM40P1** |
| BFRB9083.p1 | Female | TTM, ED | Other BFRB, TD, MDD, ANX | 12:7853028-7970710 | DEL | 117,683 | - | **SLC2A14** (1) | **Metazoa_SRP**, **NANOGP1**, **SLC2A3** |
| BFRB9094.p1 | Female | ED | MDD, ANX | 20:42605461-42650964 | DEL | 45,504 | - | **PTPRT** (1) | - |

### Exploratory gene ontology analysis

Using the subset of LoF-intolerant genes disrupted by rare CNVs in probands with BFRBs, we conducted exploratory analyses to assess the enrichment of gene ontology terms. Several gene ontology categories were significantly enriched among these eight genes, and the top enriched gene-ontology set is “regulation of synapse organization” (q-value = 0.0009) (**Supplementary Table 2**).

## Discussion

In this first parent-offspring trio genomic study of trichotillomania and excoriation disorder, our findings highlight the role of common SNPs and rare CNVs in these body-focused repetitive behavior disorders, advancing our understanding of these understudied conditions.

Our results show that probands with TTM and/or ED inherit a higher common-variant polygenic risk for OCD from their parents than expected by chance (**Figure 1.A**). This significant over-transmission of OCD polygenic risk was observed in our primary analysis of all BFRB probands of European ancestry. Our sensitivity analysis comparing BFRB probands with and without a co-occurring OCD diagnosis shows a similarly non-significant positive transmission in both groups and no significant difference between groups, highlighting that the increased transmission of OCD polygenic risk in BFRB probands may not be fully explained by the high co-occurrence of OCD with BFRBs (**Figure 1.B**). In exploratory analyses, we also found a significantly elevated transmission of OCD polygenic risk in probands from simplex families. The magnitude of the over-transmission of OCD polygenic risk was smaller and non-significant in multiplex families where a first-degree relative had TTM and/or ED (**Supplementary Figure 2**). This suggests that inheriting an elevated polygenic risk for OCD may predispose an individual to developing a BFRB, which may be especially relevant if they do not have a family history of BFRBs, though the difference between multiplex and simplex families was not statistically significant. Given the limited sample size of this study, replication of these findings in larger samples is needed.

Overall, these findings highlight potentially shared genetic factors between OCD and BFRBs. This is consistent with TTM and ED being classified as OCRDs in DSM-5 [5], the high comorbidity between BFRBs and OCD [2, 3], as well as family and twin studies that have long supported the shared liability between OCD and BFRBs [16, 17, 19, 41]. Our results also extend recent findings from Halvorson et al. [21] that demonstrate a higher burden of generalized psychiatric risk in 101 cases with TTM compared to controls. Collectively, our findings along with this past work highlight that large-scale genome-wide studies of TTM and ED may advance our understanding of the role of common genetic variants in these understudied disorders in a similar fashion to recent studies in OCD and other neuropsychiatric conditions.

In addition to common variant SNP risk, we also identified 63 rare CNVs (MAF < 0.01, length ≥ 30 kb) in probands with TTM and/or ED, with a rare CNV deletion rate of 0.32 and duplication rate of 0.4 (**Table 2**). These rates are of a similar magnitude to previously reported rates of rare CNVs identified using similar methods in other studies of TTM and OCD [21].

Among 101 individuals with TTM, Halvorsen et al. [21] identified a rare CNV (length ≥ 100 kb) deletion rate of 0.27 and a duplication rate of 0.29. In the largest rare CNV study (length ≥ 30 kb) of OCD involving 2,248 cases and 3,608 unaffected controls, Halvorsen et al. [35] report a rare deletion rate of 0.27 and a rare duplication rate of 0.32 in OCD cases compared to a deletion rate of 0.25 and a duplication rate of 0.31 in unaffected controls. We also examined the subset of large, rare CNVs (length ≥ 500 kb) in BFRB probands, with a deletion rate of 0.01 and a duplication rate of 0.03. These rates are on a similar order of magnitude to rates previously reported by McGrath et al. [36] for OCD and Tourette’s Syndrome, with the duplication rate higher than the deletion rate, in line with prior research [36]. Even though our sample size is small, our results that probands with BFRBs have similar rates of rare CNVs to those previously reported in OCRDs, and possibly higher than rates previously reported in controls, underscore that larger-scale investigations of rare CNVs in BFRBs may provide additional insight into the genetic underpinnings of these disorders in a similar manner to previous studies of related conditions.

In this study, we identified 8 rare CNVs that overlapped LoF-intolerant genes (pLI ≥ 0.9) (**Table 2**). Previous studies suggest an enrichment of rare CNVs impacting LoF-intolerant genes in OCD [36], and Halvorsen et al. [21] observed a nonsignificant difference in constrained CNVs between individuals with TTM compared to controls (OR = 3.33, *p* = 0.09). Based on this past work, we performed exploratory gene ontology analysis of the 8 LoF-intolerant genes impacted by rare CNVs in probands with BFRBs. These analyses revealed significant enrichment in several biological processes, such as synaptic organization, activity, and structure (**Supplementary Table 2**). This finding aligns with prior studies in both human and animal models that implicate the synaptic dysfunction in the pathophysiology of BFRBs [17, 42, 43], supporting the neurodevelopmental underpinnings of these disorders.

Finally, we identified 5 rare CNVs previously associated with neurodevelopmental disorders [39], with an additional 2 CNVs overlapping other neurodevelopmental risk genes [40] (**Table 3**). Of note, all the probands with these neurodevelopmental CNVs and risk genes did not have a self-reported history of intellectual disability or autism. One proband with TTM was identified to have a rare deletion in *NRXN1*, which was inherited from her mother, who has seizures. *NRXN1* encodes a presynaptic membrane cell-adhesion molecule that binds to neuroligins and is a well-established risk gene for neurodevelopmental disorders [39, 40].

Interestingly, Halvorsen et al. [21] also identified a different *NRXN1* deletion (hg38 chr2:50790067-50956421) in an individual with TTM in a cohort collected independently. In another proband with TTM, we identified a paternally inherited CNV associated with Williams-Beuren syndrome [39], a neurodevelopmental disorder characterized by distinctive facial features and mild-to-moderate intellectual disability. The proband’s CNV overlaps two Williams-Beuren syndrome-associated genes, *FKBP6* and *TRIM50* [44, 45]. In another proband, we detected two rare deletions overlapping *NIPBL* and *PTEN*, both of which are associated with neurodevelopmental disorders, and the CNV overlapping *PTEN* was identified as a CNV linked to 10q23 deletion syndrome, which is associated with developmental delays [39] (**Table 3** and **Supplementary Tables 1.2**). We also detected a deletion in a proband with TTM that has been previously associated with Prader-Willi Syndrome(PWS), which typically presents early and is characterized by developmental delay and cognitive impairment [39]. This proband’s CNV overlapped the *PWRN1* gene, which has been suggested to play a role in the etiology of PWS [46]. In a proband with ED, we identified a CNV duplication that overlaps a region associated with 8p23.1 duplication syndrome, a condition linked to learning disabilities and developmental delay [39]. Finally, one proband with ED had a duplication in kinesin-like protein *KIF1A*, which is an axonal transporter of synaptic vesicles, and has been previously associated with various neurological disorders [47], though the implications of a duplication in this gene are less well understood. Overall, these findings support a role for potentially shared genetic factors between BFRBs and neurodevelopmental conditions, consistent with BFRBs sharing stereotypic behaviors with these developmental disorders [48].

Several limitations of this study should be considered. First, ideally, control trios would be genotyped alongside cases and assessed for BFRBs to enable a more accurate comparison of burden differences in polygenic risk and CNVs. In this study, we prioritized genotyping affected parent-offspring trios and leveraged the pTDT test to compare polygenic risk in probands to their parents, which is a statistically well-powered approach to test if affected probands inherit an increased polygenic load from their parents. For rare CNVs, we compared rates in probands to those previously reported in large-scale studies of related disorders that used similar analysis methods, but future work in larger samples that simultaneously assess controls is needed to confirm the enrichment of rare CNVs in BFRBs. Second, our study examined polygenic risk for disorders commonly co-occurring with BFRBs to better understand the genomic architecture of these understudied disorders. An important limitation in the field is that there are currently no large-scale GWAS of TTM or ED, which would enable a more direct understanding of the role of common genetic variants in families impacted by BFRBs. Third, our sample had limited sex (94% female) and ancestry (95% European ancestry) diversity, and future efforts should focus on enrolling unrepresented groups to ensure our findings are generalizable. Finally, our analysis focused on common variants and rare CNVs that can be assessed by genome-wide array data, but it is likely that other variants, including rare gene-damaging single-nucleotide and indels, contribute to BFRB risk. Future large-scale DNA sequencing studies may provide further insight into the full spectrum of genetic variation that contributes to BFRB risk.

In conclusion, our study offers new insights into the genetic architecture of trichotillomania and excoriation disorder. For the first time, we reported a significant over-transmission of polygenic risk for OCD in probands with these body-focused repetitive behavior disorders. We also identified several rare CNVs that overlap LoF-intolerant and neurodevelopmental risk genes. These findings represent an important step forward toward understanding the genomic underpinnings of BFRBs. Future efforts leveraging larger BFRB cohorts may continue to deepen our understanding of trichotillomania and excoriation disorder genomics and pathophysiology.

## Data Availability

All data produced in the present study are available upon reasonable request to the authors.

## Acknowledgements

We would like to thank the families who contributed to this research study, as well as the TLC Foundation for BFRBs for their support of our recruitment efforts. We would also like to acknowledge research assistants Kayla Delapenha and Jessica Levine, who contributed to the recruitment and collection of data. This research was supported by the National Institutes of Mental Health of the National Institutes of Health [under award numbers K08MH128665 to E.O., R01MH114927 to T.V.F.], the American Academy of Child and Adolescent Psychiatry (E.O.), and the Alan B. Slifka Foundation Riva Ariella Ritvo endowment (E.O.). S.R.G. was supported by CTSA Grant Number TL1 TR001864 from the National Center for Advancing Translational Science of the NIH. S.B.A. was supported by training grants from the National Institutes of Mental Health of the National Institutes of Health [T32 MH18268 and R25 MH077823]. I.M. was supported by the Interdepartmental Neuroscience Program NIH T32 Training Grant, 5T32NS041228-24. The content is solely the responsibility of the authors and does not necessarily represent the official views of the National Institutes of Health.

