## Supplementary information for "A genomic study of trichotillomania and excoriation disorder in families"

File Includes:

Supplementary Tables 1-2
Supplementary Figures 1-4
Supplementary Methods
Supplementary Discussion
Supplementary References

**Supplementary Table 1.1.** CNV predefined by Kendall et al. (2019) as neurodevelopmental (54 CNVs total)

| CNV locus in BFRB proband (hg38) | CNV locus in Kendall (hg38) | CNV type | Syndrome |
|---|---|---|---|
| chr10:87894024-87925562 | chr10-80285716-87171894 | DEL | 10q23 |
| chr15:24140203-24249920 | chr15:23067755-28145193 | DEL | PWS/AS |
| chr2:50652887-50725225 | chr2-49918505-51032536 | DEL | *NRXN1* |
| chr7:73308984-73391588 | chr7-73330912-74728554 | DUP | Williams–Beuren syndrome |
| chr8:8243957-8465532 | chr8-8241468-12015049 | DUP | 8p23.1 |

Copy number variants (CNVs) identified in probands with BFRBs that overlapped CNVs previously associated with neurodevelopmental disorders defined by Kendall et al. [3] (lifted over to hg38). CNV, copy number variant; DEL, deletion; DUP, duplication; PWS/AS, Prader-Willi Syndrome/Angelman Syndrome.

**Supplementary Table 1.2.** Gene association results from Fu et al. (2022) for Autism Spectrum Disorder, Developmental Delay, and Neurodevelopmental Disorders

| Gene | FDR_TADA_ASD | FDR_TADA_DD | FDR_TADA_NDD |
|---|---|---|---|
| *KIF1A* | 0.823772021 | 0 | 0 |
| *NRNX1* | 9.95963E-14 | 0.44196728 | 9.85689E-15 |
| *NIPBL* | 9.95403E-06 | 0.014825475 | 2.82718E-08 |
| *PTEN* | 0 | 0 | 0 |

Copy number variants (CNVs) identified in probands with BFRBs that overlapped CNVs harboring neurodevelopmental risk genes with an FDR ≤ 0.05 in Fu et al [4]. FDR_TADA_ASD, TADA Autism Spectrum Disorder False Discovery Rate of the gene; FDR_TADA_DD, TADA, developmental delay false discovery rate of the gene; FDR_TADA_NDD, TADA, neurodevelopmental disorders false discovery rate of the gene.

**Supplementary Table 2.** Gene-ontology-based sets that are enriched for 8 genes harboring rare, large damaging variants in BFRB probands

| Gene ontology term | Category, level | Set size | Genes contained | p-value | q-value |
|---|---|---|---|---|---|
| GO:0050807 regulation of synapse organization | BP5 | 210 | 3 | 4.16E-05 | 1E-03 |
| GO:0050803 regulation of synapse structure or activity | BP3 | 220 | 3 | 4.78E-05 | 0.002 |
| GO:0050808 synapse organization | BP3 | 410 | 3 | 0.0003 | 0.007 |
| GO:0051128 regulation of cellular component organization | BP4 | 2562 | 5 | 0.0006 | 0.034 |
| GO:0099173 post synapse organization | BP3 | 160 | 2 | 0.0013 | 0.02 |
| GO:0034613 cellular protein localization | BP4 | 1927 | 4 | 0.0025 | 0.068 |
| GO:0070727 cellular macromolecule localization | BP3 | 1937 | 4 | 0.0026 | 0.029 |
| GO:0051960 regulation of nervous system development | BP5 | 894 | 3 | 0.0029 | 0.035 |
| GO:0008017 microtubule binding | MF5 | 271 | 2 | 0.0038 | 0.004 |
| GO:0008092 cytoskeletal protein binding | MF3 | 989 | 3 | 0.003 | 0.027 |
| GO:0006928 movement of cell or subcellular component | BP2 | 2164 | 4 | 0.0039 | 0.113 |
| GO:0050890 cognition | BP4 | 277 | 2 | 0.0040 | 0.071 |
| GO:0016791 phosphatase activity | MF5 | 286 | 2 | 0.0042 | 0.004 |
| GO:0051348 negative regulation of transferase activity | BP5 | 288 | 2 | 0.0043 | 0.035 |
| GO:0007399 nervous system development | BP4 | 2351 | 4 | 0.0054 | 0.072 |
| GO:0097060 synaptic membrane | CC2 | 370 | 2 | 0.0070 | 0.08 |
| GO:0015631 tubulin binding | MF4 | 372 | 2 | 0.0071 | 0.011 |
| GO:0042578 phosphoric ester hydrolase activity | MF4 | 379 | 2 | 0.0074 | 0.011 |
| GO:0051130 positive regulation of cellular component organization | BP5 | 1270 | 3 | 0.0078 | 0.044 |
| GO:0008104 protein localization | BP3 | 2640 | 4 | 0.0082 | 0.073 |

Gene ontology terms that were enriched among genes that were intolerant to loss-of-function variation that overlapped rare CNVs identified in BFRB probands. Gene ontology, GO; BP, biological process; MF, molecular function; CC, Cellular Component.

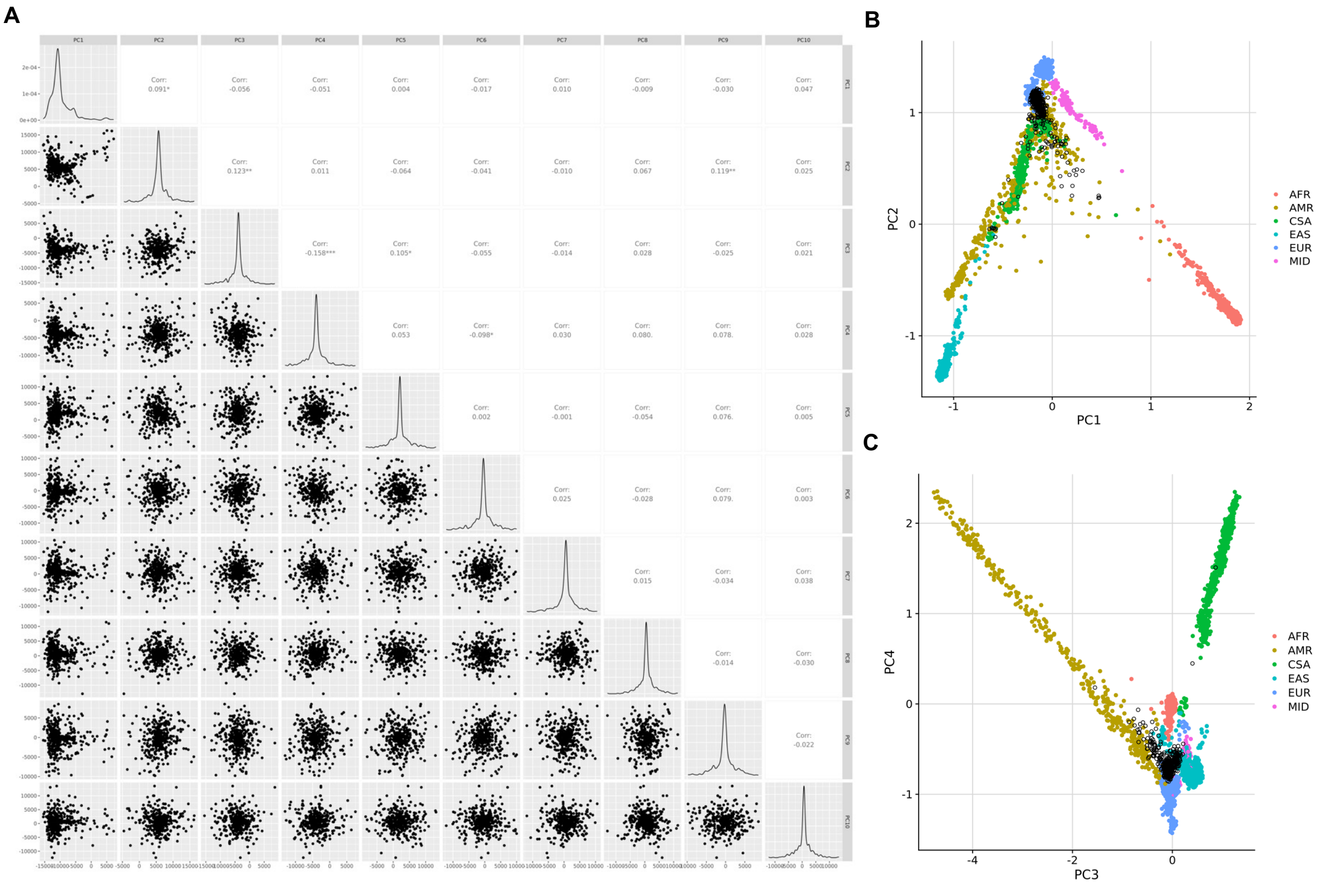


**Supplementary Figure 1.** Plots from principal component analysis to identify outliers based on genome-wide array metrics. **A**) Shows the percentage of variance captured by the first 10 principal components from the genome-wide array data from cases and controls. **B**) Show the first four principal components based on the PCA that captures ancestry clusters. This figure includes PCA outliers (>5 standard deviations in PCAs 1-3) removed during quality control. The first PC is the only one with a detectable difference (logistic regression $p < 0.01$) between samples and was used as a covariate in subsequent GWAS and PRS analyses. AFR = African; AMR = Admixed American; EAS = East Asian; EUR = European; CSA = Central and South Asian; MID = Middle Eastern.

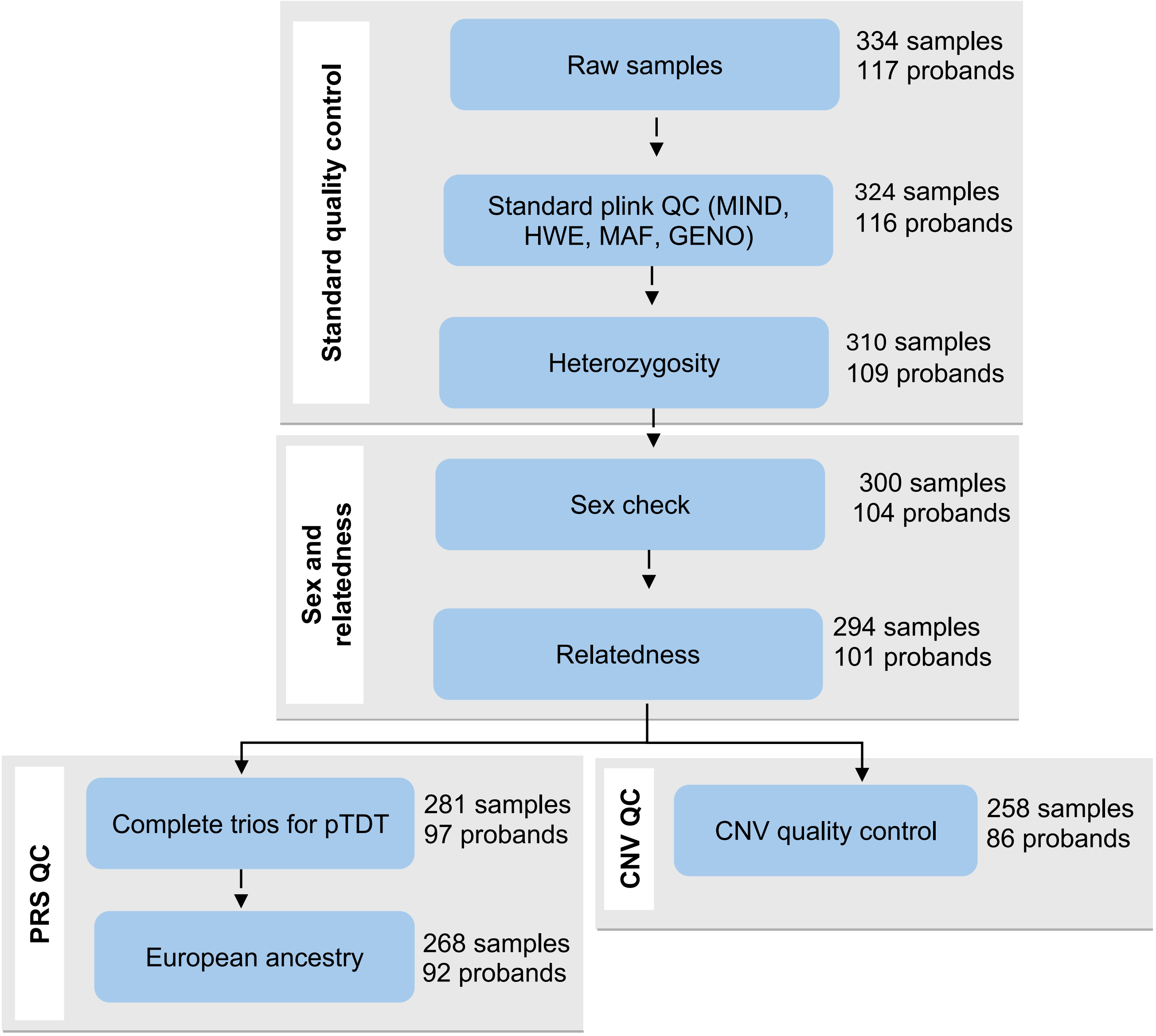


**Supplementary Figure 2.** Sample-level quality control workflow of genome-wide array data. Workflow outlines the quality control (QC) steps applied to the raw BFRB genome-wide array data, beginning with standard QC metrics including missingness (MIND), Hardy-Weinberg Equilibrium Fisher's (HWE), and minor allele frequency (MAF), followed by heterozygosity filtering. Samples were then filtered if discrepancies were detected between self-reported gender and sex chromosome data or relatedness. Subsequent QC steps were applied separately for polygenic risk score (PRS) and copy number variant (CNV) analyses. For PRS QC, only complete trios and quads of confirmed European ancestry were retained. For CNV analysis, additional sample-level quality control was performed using PennCNV's filter_cnv.pl tool, which identifies low-quality samples based on parameters such as waviness factor (|WF|), B-allele frequency (BAF) drift, and LRR standard deviation (LRR_SD). The number of samples and probands retained after each step is shown. MIND, sample missingness; HWE, Hardy-

Weinberg equilibrium; MAF, minor allele frequency; GENO, genotype-call missingness; |WF|, waviness factor; BAF, B-allele frequency; LRR_SD, Log R Ratio standard deviation.

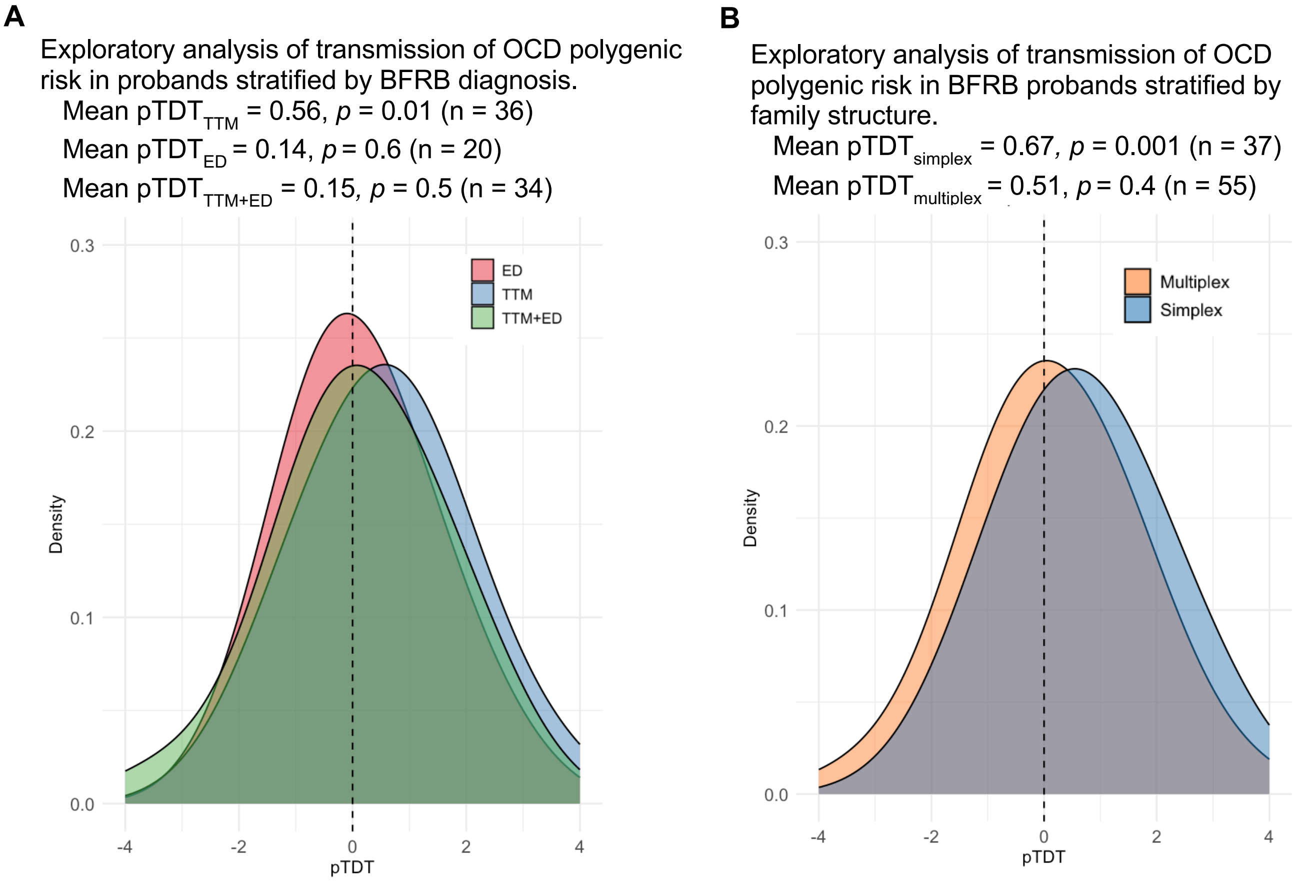


**Supplementary Figure 3. Exploratory stratified analyses exploring impacts of BFRB diagnosis and family structure on transmission of obsessive-compulsive disorder (OCD) polygenic risk in probands of European Ancestry with trichotillomania (TTM) and/or excoriation disorder (ED). A)** Probands with a singular diagnosis of TTM, ED, and TTM+ED all show an OCD mean pTDT in the positive direction with only the TTM reaching statistical significance (mean $pTDT_{TTM}$ = 0.56, $p$ = 0.01, n = 36; mean $pTDT_{ED}$ = 0.14, $p$ = 0.6, n = 20; mean $pTDT_{TTM+ED}$ = 0.15, $p$ = 0.5, n = 34). The mean difference in pTDT between these body-focused repetitive behavior (BFRB) groups did not reach statistical significance ($p$ = 0.3). **B)** Simplex trios show a significant over-transmission of OCD polygenic risk (mean pTDT = 0.67, $p$ = 0.001; n = 37), while multiplex trios do not show a significant increased transmission (mean pTDT = 0.15, $p$ = 0.4; n = 55). The mean difference in pTDT between simplex and multiplex trios was not statistically significant ($p$ = 0.06).

**Supplementary Figure 4.** Plots of Log R Ratios and B Allele frequencies for the 11 rare copy number variants (CNVs) identified in BFRB probands that are described in Table 3. Plots are presented for the proband and both of their parents. All CNVs were validated based on the intersection of the call sets from both PennCNV and CnvPartition, as described in the methods and supplemental methods. Plots were generated from aligned; normalized genome-wide array reads from Genome Studio.

**A.** Chr2:240684041-240762707 (Duplication) for BFRB9003.p1, BFRB9003.mo, and BFRB9003.fa

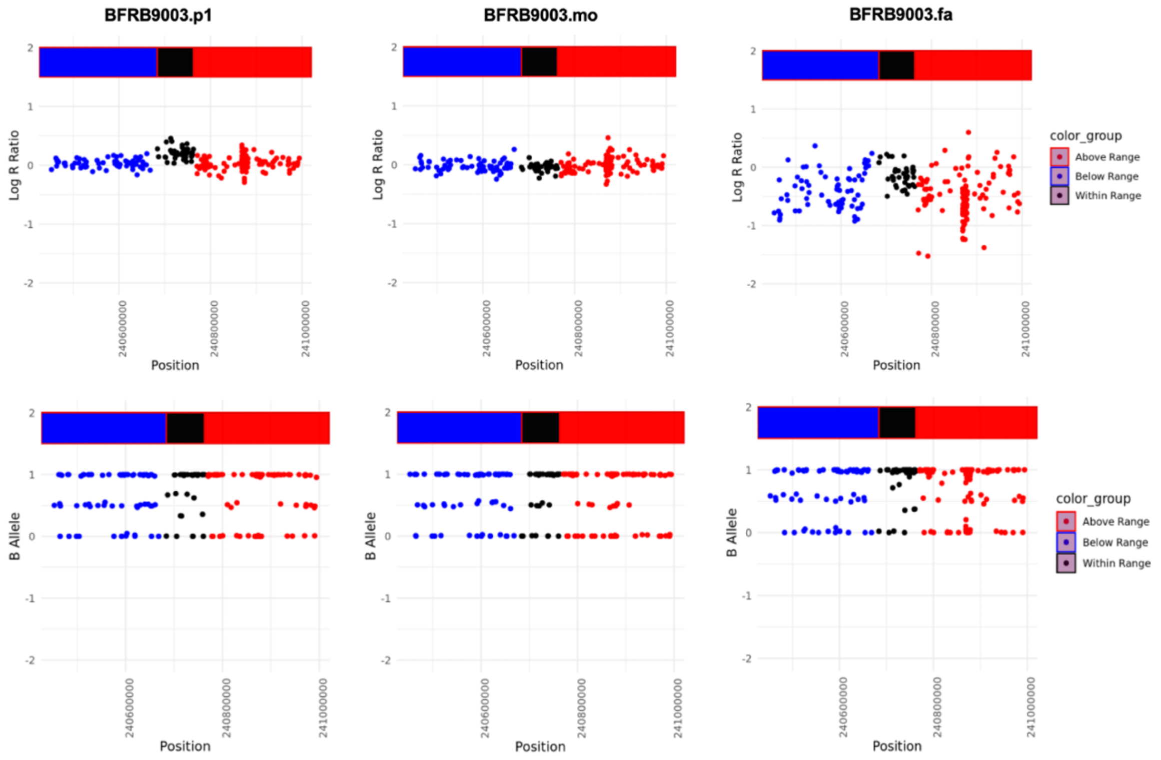

**B.** Chr8:73040513-73159092 (Duplication) for BFRB9003.p1, BFRB9003.mo, and BFRB9003.fa

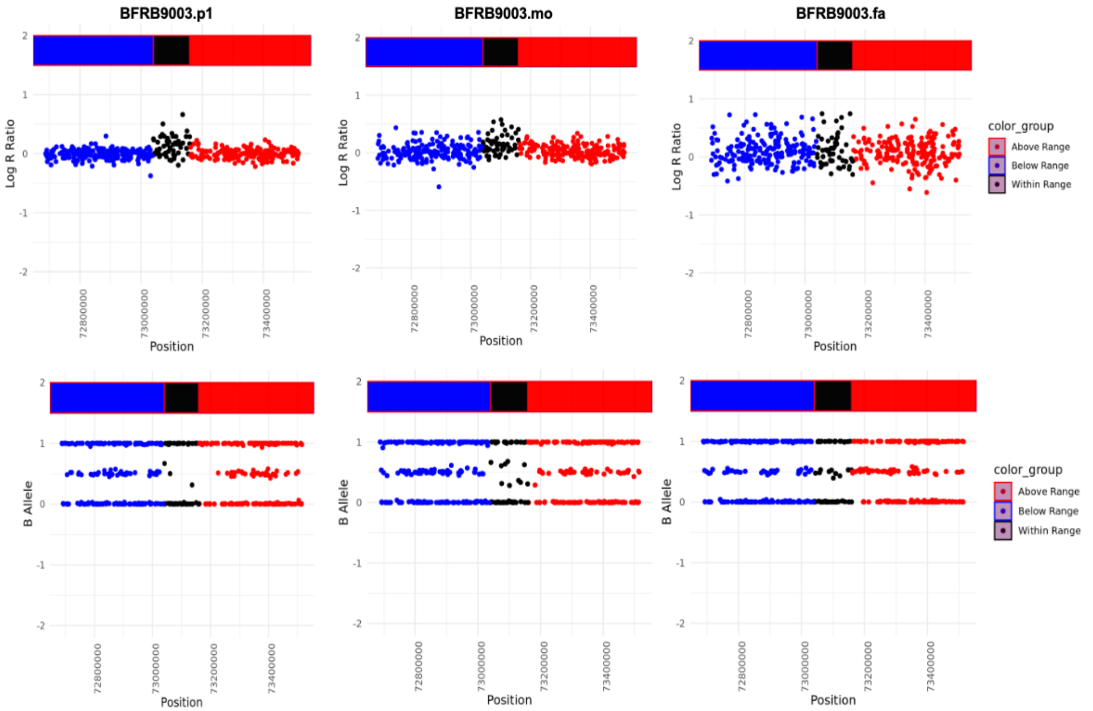


**C.** Chr2:50652887-50725225 (Deletion) for BFRB9021.p1, BFRB9021.mo, and BFRB9021.fa

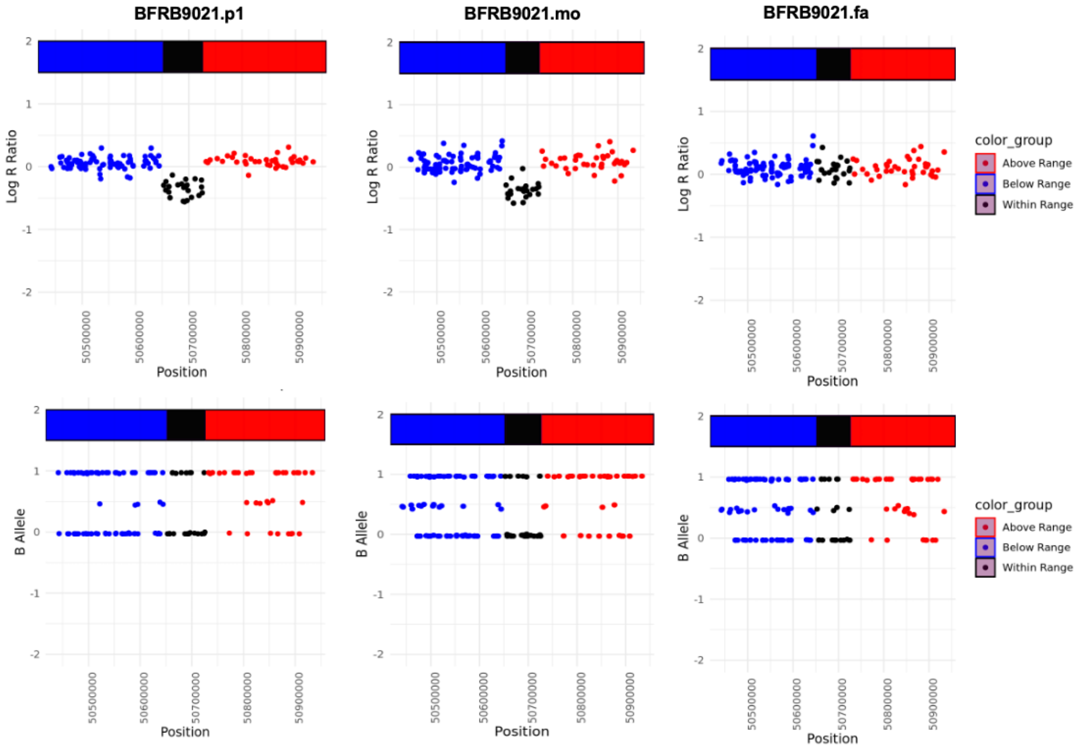

**D.** Chr7:73308984-73391588 (Duplication) for BFRB9023.p1, BFRB9023.mo, and BFRB9023.fa

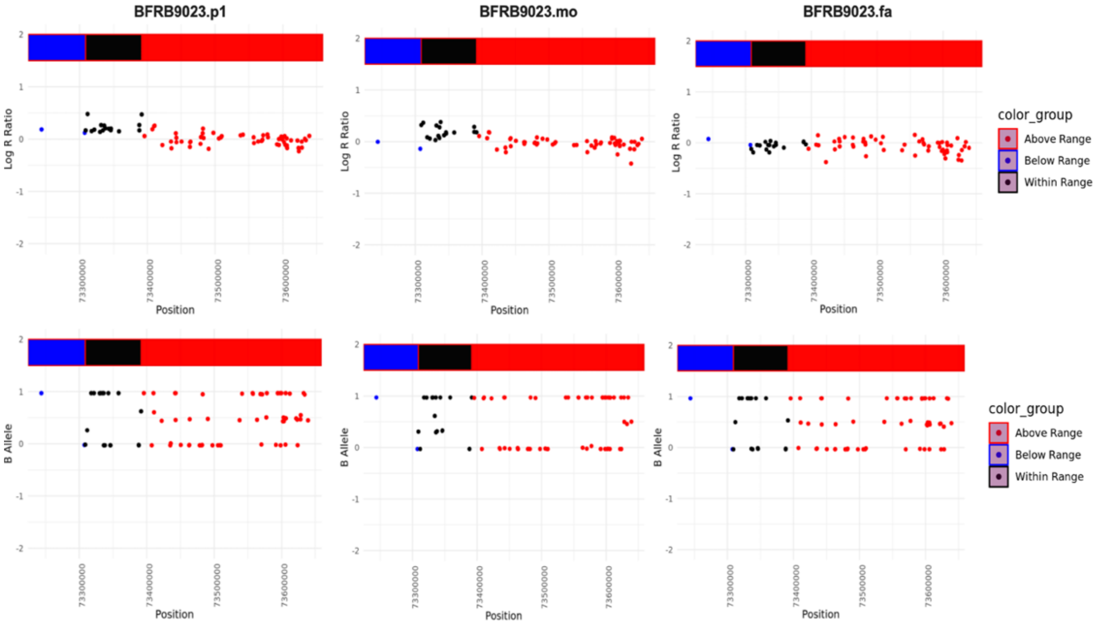


**E.** Chr5:37000986-37046181 (Deletion) for BFRB9047.p1, BFRB9047.mo, and BFRB9047.fa

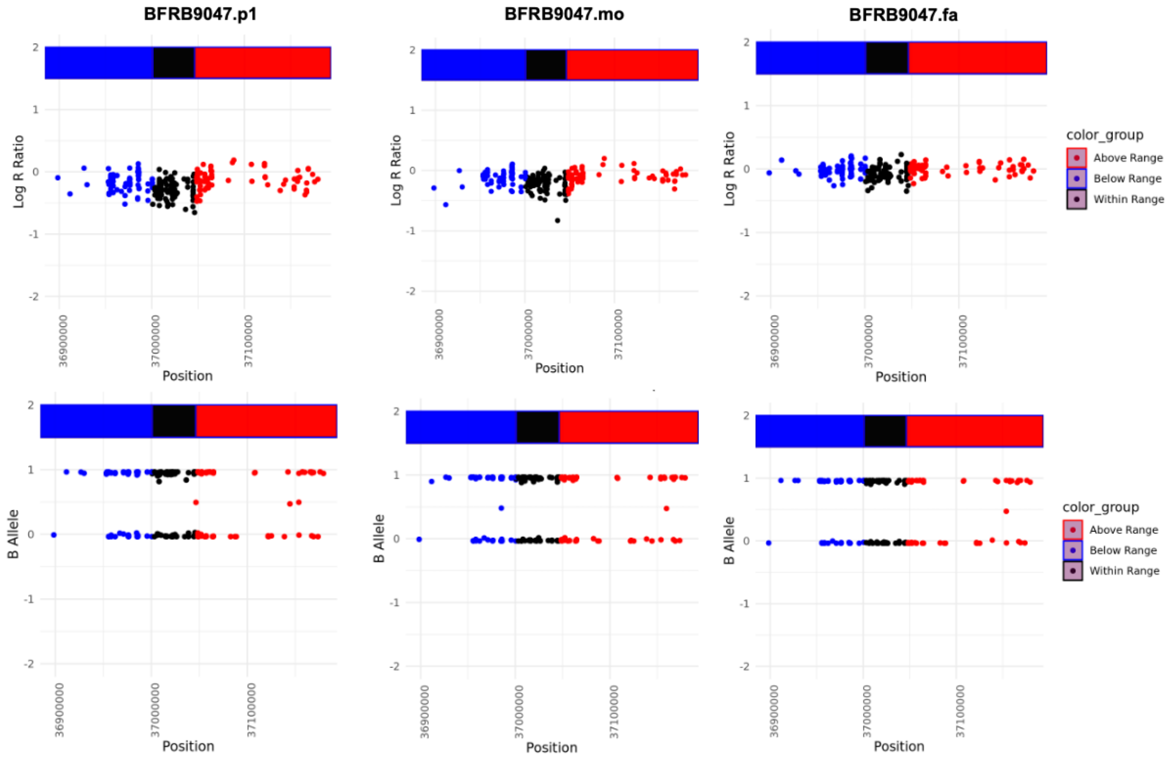

**F.** Chr10:87894024-87925562 (Deletion) for BFRB9047.p1, BFRB9047.mo, and BFRB9047.fa

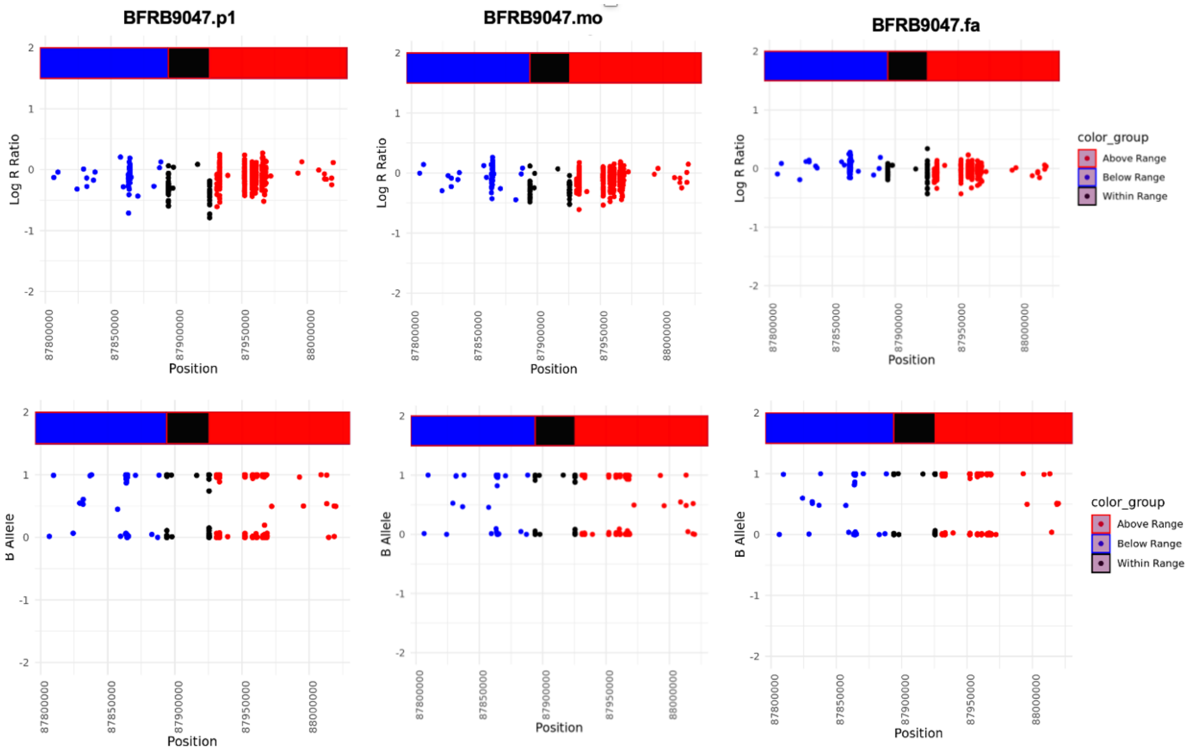

**G.** Chr9:101144710-101244151 (Deletion) for BFRB9059.p1, BFRB9059.mo, and BFRB9059.fa

**H.** Chr15:24140203-24249920 (Deletion) for BFRB9061.p1, BFRB9061.mo, and BFRB9061.fa

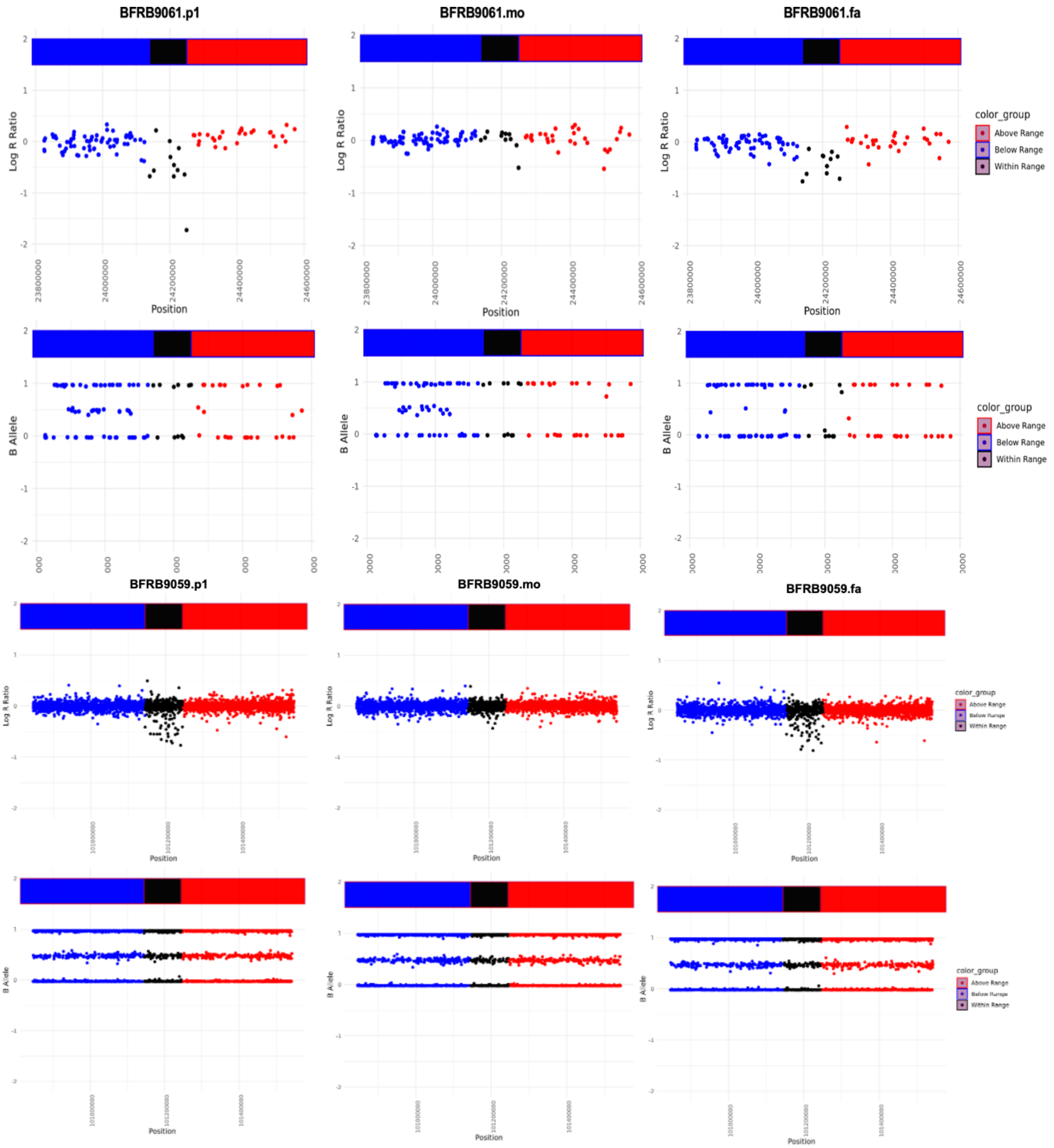

**I.** Chr8:8243957-8465532 (Duplication) for BFRB9072.p1, BFRB9072.mo, and BFRB9072.fa

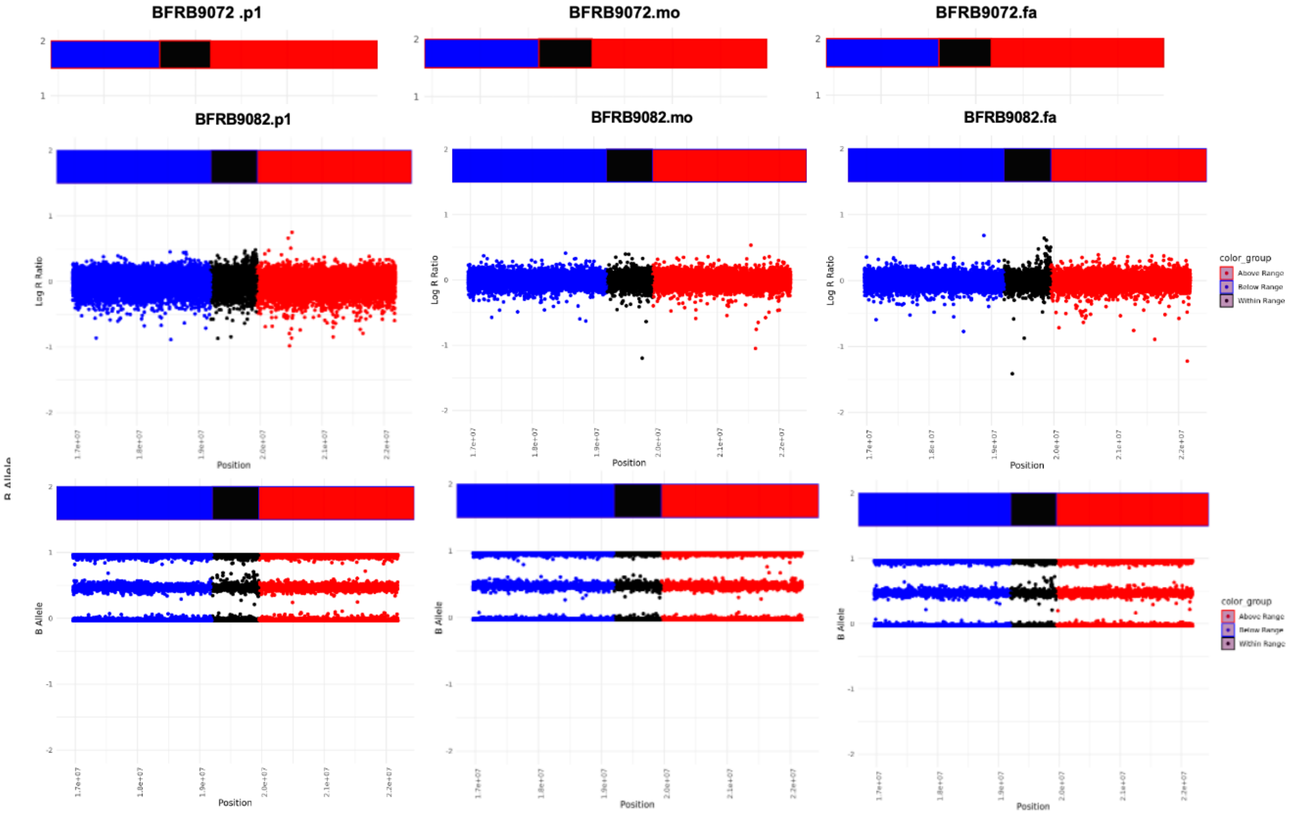


**J.** Chr14:8243957-8465532 (Duplication) for BFRB9082.p1, BFRB9082.mo, and BFRB9082.fa

**K.** Chr12:7853028-7970710 (Deletion) for BFRB9083.p1, BFRB9083.mo, and BFRB9083.fa

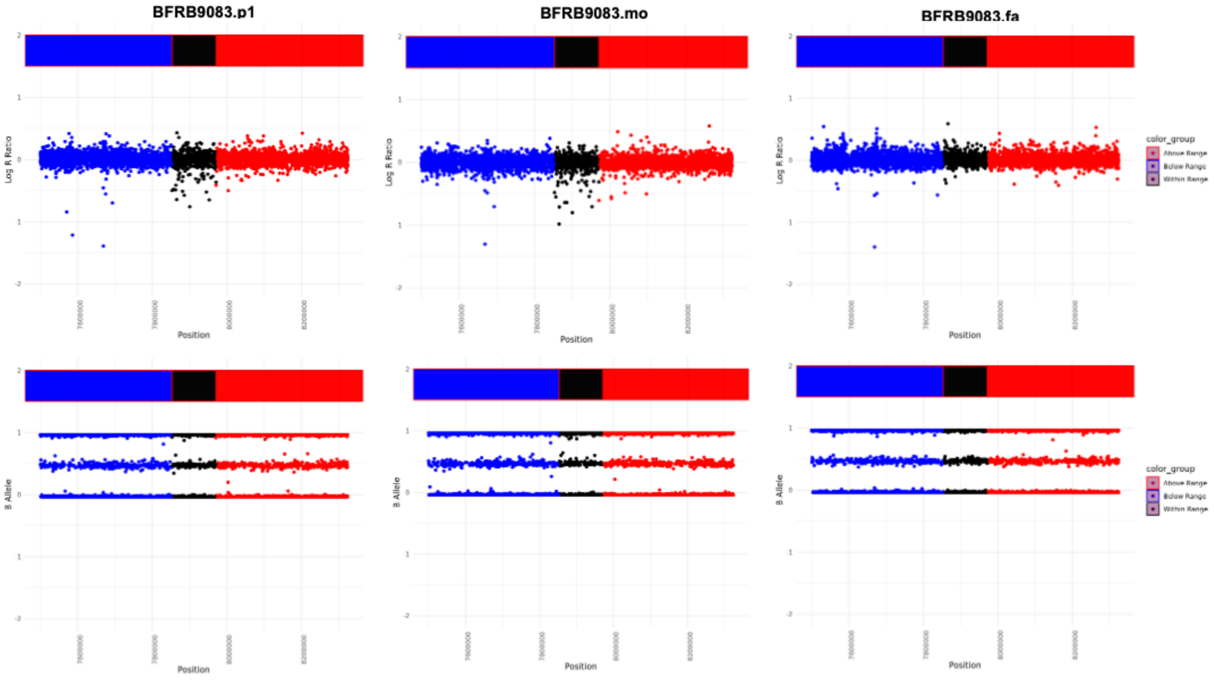


**L.** Chr20:42605461-42650964 (Deletion) for BFRB9094.p1, BFRB9094.mo, and BFRB9094.fa

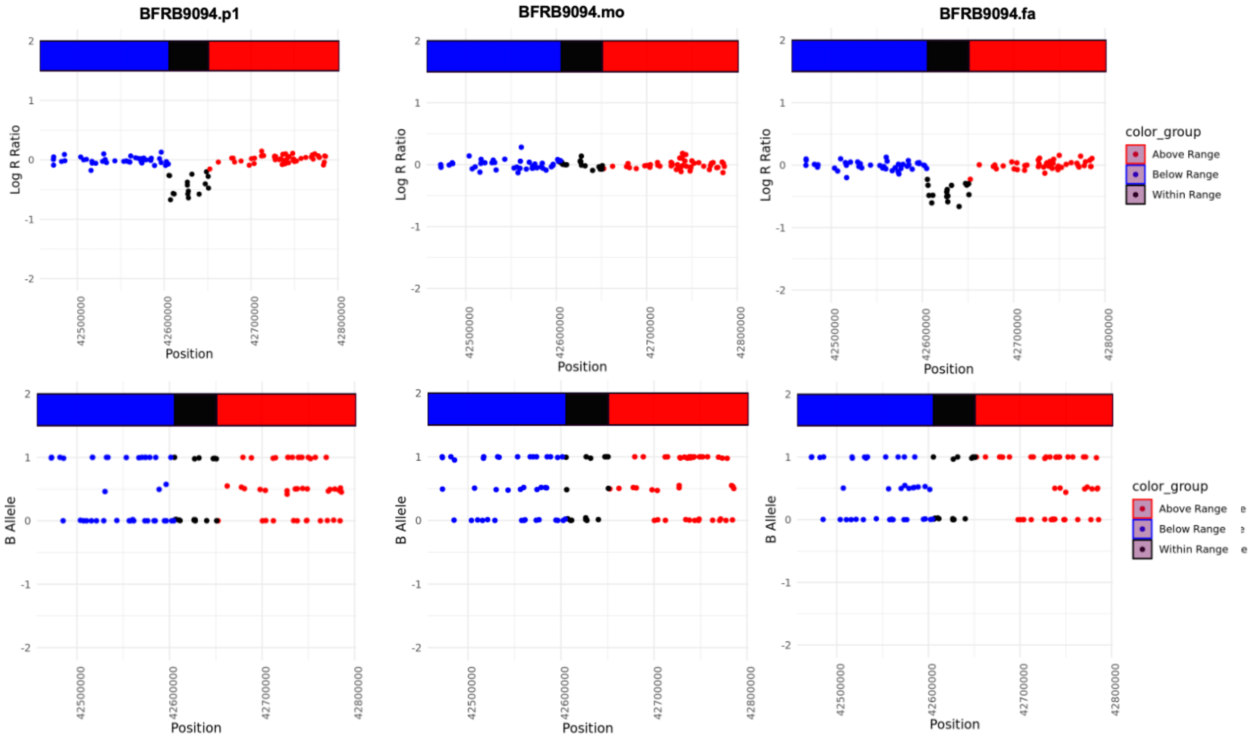

### Supplementary Methods

#### *Ancestry matching*

We used GenoPred (opain.github.io/GenoPred) to perform principal component (PC) analysis on the BFRB dataset using reference data from the 1000 Genomes Phase 3 (1KG) and Human Genome Diversity Project panels [1]. GenoPred calculates the first six principal components and projects them onto the target dataset. Ancestry inference was then performed using the first four PCs, matching everyone to an ancestry group based on the reference panels: AFR (African), AMR (Admixed American), EAS (East Asian), EUR (European), CSA (Central and South Asian), and MID (Middle Eastern). In this sample, 92/97 complete trios who passed QC were of European ancestry, which was used in the primary analyses.

#### *PennCNV and cnvPartition*

Standard Illumina cluster files were used to generate LogR ratios (LRR) and B Allele Frequencies (BAF) in Illumina GenomeStudio. CNV calls were generated using PennCNV (version 2010-05-01) and cnvPartition version 3.2.0 (http://www.illumina.com/software/illumina_connect.ilmn).

PennCNV uses an integrated hidden Markov model that incorporates multiple sources of information, including total signal intensity and allelic intensity ratio at each SNP marker, the distance between neighboring SNPs, the allele frequency of SNPs, and pedigree information (http://www.openbioinformatics.org/penncnv/).

CnvPartition uses a likelihood-based method that estimates copy number by comparing the observed log R ratio (LRR) and B allele frequency (BAF) for each locus. CnvPartition also provides a CNV confidence score when a CNV is detected, which is defined as the sum of all logged likelihoods in the region for the assigned copy number minus the sum of all log L2 values for loci in the region.

#### *Merging of PennCNV and CnvPartition Calls*

PennCNV and cnvPartition call sets were merged using in-house scripts leveraging the GRanges package from Bioconductor [2]. CNV data from both call sets were first parsed to extract the chromosome, start, and end coordinates of the CNV, then converted into genomic ranges. We then identified overlapping CNV regions between the two call sets. Only CNVs with an overlap greater than 50% of their length were retained to ensure high-confidence matches.

#### *CNV visualization*

We examined all rare copy number variants of interest entering our analyses (**Table 3**) by visualizing reads from genome-wide array data using in-house scripts for the proband, father, and mother (**Supplementary Figure 4**). These scripts visualize the area of the CNV and provide the log R ratio and B-allele frequency using the normalized, aligned genome-wide array data to confirm deletion or duplication.

#### *Exploratory pathway analysis*

For the exploratory gene set over-representation analysis, we used the following default settings in ConsensusPathDB: a minimum overlap of 2 genes with the input list, a p-value cutoff of 0.01, and gene ontology categories from levels 2 to 5. Fisher's exact tests are used to calculate p-values within each database, which are corrected for multiple comparisons. ConsensusPathDB also calculates q-values, which are adjusted for the number of tests performed across all databases. Q-values are derived using the Benjamin–Hochberg (BH) correction method based on the p-values from Fisher's exact test. Results can be found in **Supplementary Table 2**.
